# Augmenting Fact and Date of Death in Electronic Health Records using Internet Media Sources: A Validation Study from Two Large Healthcare Systems

**DOI:** 10.1101/2025.01.24.25321042

**Authors:** Michele LeNoue-Newton, Mohammed al-Garadi, Kerry Ngan, Haritha Pillai, Ruth M. Reeves, Daniel Park, Dax M. Westerman, José J. Hernández-Muñoz, Xi Wang, Aida Kuzucan, Shirley V. Wang, Kueiyu Joshua Lin, Candace Fuller, Melissa McPheeters, Michael E. Matheny, Rishi J. Desai

**Affiliations:** Department of Biomedical Informatics, Vanderbilt University Medical Center, Nashville, TN, USA; Division of Pharmacoepidemiology and Pharmacoeconomics, Brigham and Women’s Hospital and Harvard Medical School, Boston, MA, USA; Office of Surveillance and Epidemiology, Center for Drug Evaluation and Research, US Food and Drug Administration, Silver Spring, MD; Department of Population Medicine, Harvard Medical School and Harvard Pilgrim Health Care Institute, Boston, MA, USA; RTI International, Research Triangle Park, NC, USA; Geriatrics Research Education and Clinical Care Service, Tennessee Valley Healthcare System VA, Nashville, TN, USA

**Author notes:** **Correspondence:** Rishi J Desai, PhD, Associate Professor of Medicine, Division of Pharmacoepidemiology and Pharmacoeconomics, Department of Medicine, Brigham and Women’s Hospital, Harvard Medical School, 1620 Tremont Street, Suite 3030-R, Boston, MA 02120, USA, | |.

**Keywords:** vital statistics, mortality, online media, data linkage

## Abstract

**Objective:** To evaluate the validity of death ascertainment from publicly available internet media (IM) sources by benchmarking against state and Federal vital statics data for patients in two large healthcare systems from the US.

**Methods:** We extracted names and dates of birth and death from publicly available data—including obituaries and memorial websites—using previously developed natural language processing models. These data were probabilistically matched to electronic health records (EHRs) from Mass General Brigham (MGB) and Vanderbilt University Medical Center (VUMC) on first name, last name, and date of birth. Using reference standards from state vital statistics databases from MA, CT, and VT for MGB and the National Death Index (NDI) for VUMC patients, we reported positive predicted values (PPV) considering cases where dates of death from IM sources were within 7 days of the reference standard to be true positives. We also reported sensitivity of deaths ascertained from IM sources.

**Results:** When probabilistically matching 8.1 million deaths extracted from public data to 78,848 deaths observed in the reference standards across two sites, 30,607 (38.8%) matched exactly. A PPV of 98.2% for MGB and 98.9% for VUMC was observed for exact matches, while <6% for non-exact matches. Considering only the exact matches, IM sources led to an improvement in sensitivity of death capture by 24% in MGB and 18% in VUMC, compared to using EHRs alone for death ascertainment.

**Conclusions:** Using public information to augment mortality data increased capture of death meaningfully over reliance on EHR records alone.

## 1.0 Background

In safety and effectiveness evaluations of medications, mortality is often an important outcome of interest to researchers, clinicians, and patients. However, information regarding deaths that occur outside of a clinical setting are frequently missing in real-world data sources such as electronic health records (EHRs) or insurance claims^1-3^. Under-ascertainment of mortality can lead to bias in studies aimed at evaluating effectiveness or harms of given treatments or conditions^3-8^. Thus, substantial effort has gone into identifying alternate sources for death data to augment information available in claims or EHRs, by linking these data to external sources of information^3, 9-11^.

In the United States, deaths are tracked at the state and national levels through vital statistics registries^12, 13^. Reporting into electronic death registries occurs initially at the state-level usually within 24 to 48 hours with data then available for public use 1-2 years from the date of collection. Currently, data are not routinely linked between state death registries and EHRs or claims data. Several all-cause death databases exist, including the Social Security Death Index (SSDI), CenSoc^14, 15^, the National Death Index (NDI), Veteran Administration data, and Center for Medicare Services (CMS) data^16^. Each of these databases have specific biases, delays in release, or are not readily available to researchers. CenSoc and SSDI data skew towards older populations and only contain individuals with social security numbers (SSN), which can lead to bias in racial-ethnic distributions^14, 15, 17^. Missingness in SSDI has been exacerbated by regulatory changes made to the national SSDI in 2011 that prevent significant portions of state vital statistics data from being included^18, 19^. NDI remains a highly accurate source of information but has significant time delays and is prohibitively expensive for large scale research studies and has some bias in linkage towards individuals with SSNs^20, 21^.

Internet based resources have been used for public health surveillance, including pharmacovigilance studies of adverse events, but generally on a population rather than an individual level^22-27^. There are several publicly available data sources that could potentially be leveraged to augment death ascertainment. While cause of death information is largely unavailable outside of narrative notes, death certificates, and autopsy reports, the date of death has the potential to be detected through several mechanisms. Obituaries and memorial websites can offer near real-time updates on vital status^26, 28-30^, especially if posted electronically where they can be easily accessed and processed using natural language processing (NLP) tools. Other sources of information on social media may also provide indications that an individual has died, including GoFundMe and similar sites on which family members or friends may share information.

Several prior studies have used on-line obituary data to extract fact and date of death;^10, 11, 29-31^ however, all of these studies were focused on cohorts of patients with specific diagnosis, for instance cancer or scleroderma. These prior studies also did not explore sources such as memorial websites and other social media in addition to obituaries. To that end, we aimed to provide health system level validation of death information retrieved using previously developed NLP models from various online sources. We linked NLP derived death information to EHRs from two large tertiary care medical centers using probabilistic linkage methodology and benchmarked against >78,000 deaths identified from state and Federal vital statistics data as the gold standard information.

## 2.0 Methods

### 2.1 Data Sources

#### 2.1.1 Internet Media (IM) Data

Decedent names, birth dates, and dates of death were extracted from publicly available IM posts from Obituary.com, GoFundMe, and Everloved/TributeArchives. These data were compiled into a database to facilitate linkage with patient populations for analysis by both MGB and VUMC (Figure 1A). Data was extracted either from structured metadata or by using an XGBoost transformer model trained at VUMC based on manual annotations of 3150 documents (1050 from each source) by trained nurse annotators. Details on the training and models for extraction of mortality data from IM are described elsewhere^32^. The majority of the 8M+ IM records were derived from Obituary.com (91%) with some records from Everloved/TributeArchives (9%), and a small amount from GoFundMe (<1%) (Figure 1B). All records in the final IM cohort were required to have complete entries for first name, last name, date of birth, and date of death, with no missing data.

#### 2.1.2 MGB Reference Cohort

The MGB reference cohort consisted of 65,139 deceased individuals from an MGB EHR-Medicare claims linked data asset. All of these deaths were recorded between Jan 1, 2015, and Dec 31, 2020, in one of 3 state vital statistic database (MA, CT, VT) and included patients who had ≥1 visit recorded in MGB EHRs within 180 days prior to death.

#### 2.1.3 VUMC Reference Cohort

As it was not feasible to submit all VUMC patients within the eligibility cohort of individuals who had either an inpatient or outpatient encounter at VUMC between Oct 1, 2015 and Jan 1, 2020 (over 1M patients) to search for deaths records in NDI due to cost consideration, we submitted a subsample to NDI to obtain a “gold-standard” reference for deaths. Patients were initially stratified by availability of a recorded death date and year of last known contact in accordance with NDI submission protocols. We limited submission to those with last known contact dates or recorded death dates between Jan 1, 2019, and Dec 31, 2021, and to patients who had ≥1 visit in the year prior to the last date of contact recorded at VUMC. Over 300,000 patients met these criteria, 29,233 patients with recorded deaths and 278,576 patients with unknown vital status, from these we selected a subsample of 35,015 individuals. Patients with recorded deaths were oversampled to ensure adequate sample sizes for related studies using the cohort related to cause of death.

Of the 35,015 patients submitted, 12,395 patients had a death recorded within VUMC records, while the remaining 22,620 patients had an unknown vital status. Patients with unknown vital status were selected roughly equally from across the 3 submission years with approximately 7500 selected per year. Patients were selected randomly from each stratification grouping. NDI provided potential matches from the national registry along with match characteristics. We selected the highest probability match for each patient for assessment. We grouped potential matches based on the data elements matched (SSN, first name, last name, sex, and date of birth) and evaluated each group on concordance with recorded dates of death to establish deterministic match criteria that would result in the highest precision and recall. The following criteria were used to determine the final match cohort. Matches with a full SSN match and mismatches on a combination of two of the following elements: first name, last name, day of birth, month of birth, or year of birth. If DOB was the only mismatch, all 3 elements could be mismatched given that year of birth was within 10 years. If SSN was not a match, then all the following elements needed to match exactly: first name, last name and DOB. Determination of the final matched cohort aligned criteria between institutions and established similar criteria for matching between NDI and EHR as used for IM to EHR.

### 2.2 Data Linkage and Validation

#### 2.2.1 Linkage of IM Records to EHR Records

Both MGB and VUMC utilized SPLINK^33^ to determine probabilistic matches between IM records and their reference cohort. First name, last name, date of birth, and state of residence were used to match patient records between sources. The algorithm was trained independently at each site with the full IM dataset and each institution’s reference cohort using identical parameters. The training determined weights for each variable and allowed for calculation of matching weights and probabilities for all matches within the dataset. Initial threshold for potential matching was set as a probability of 0.80 and the highest probability match for each reference patient was selected as the best match for further analysis.

#### 2.2.2 Validating Linkage of IM Records to EHR Records

To determine the probability cut-off that yielded the matches with highest accuracy, we manually reviewed the potential matches identified using the probabilistic matching algorithm for validity using stratified random sampling of three hundred matches grouped by probability scores separately at each institution. Accuracy for the probabilistic matching algorithm was defined as the proportion of records predicted by the algorithm as matches that were confirmed by manual reviewers. For the initial round of validation, we limited data to data elements available to the match algorithm and assessed whether the match was potentially the correct patient. The following criteria were used in the manual review for determination of a potential match: 1) Exact first name with DOB within 1-5 days, 2) Exact match on uncommon first name and last name with different DOB (e.g. common names like John Smith would not match if DOB was not a match), 3) manual reviewers were free to use their discretion to account for nicknames (e.g. Sue instead of Susan), typos, and misspellings.

Positive predicted values (PPVs) were calculated based on alignment of date of death derived from the IM sources and the date of death from the reference standard for each match. For these calculations, we considered cases where dates of death from IM sources were within 7 days of the reference standard to be true positive cases. We also reported results based on exact match of date of death with the reference standard.

### 2.3 Ascertainment of Augmentation

Augmentation of date of death data from IM sources was determined at MGB by reporting the sensitivity of capture of deaths from IM records and EHRs as a proportion of total deaths identified in the reference cohorts. To calculate comparable metrics at VUMC, we adjusted for the oversampling of known deaths in the NDI-matched reference cohort. To estimate the actual impacts of IM sources on augmentation of death data, we first determined the percentage of patients with unknown death status submitted to NDI where a matched death record was identified to estimate the number of patients within the VUMC EHR that have unrecorded deaths. We then calculated the percentage of our reference cohort that had deaths identified within IM sources but not recorded in the EHR to estimate the number of deaths that would be identified if all patients were matched to public online data.

## 3.0 Results

### 3.1 NDI Match Results for patients sampled from the VUMC Research Derivative database

The final VUMC cohort contained 13,709 deaths identified in the NDI “gold-standard” reference. Of these, 11,934 had a death recorded in the EHR and 1,775 deaths were identified within the unknown cohort of 22,620 submitted to NDI (Table 1). This translates to 7.8% of persons with unknown death status matching with a deceased record.

**Table 1:** Stratification of patients meeting inclusion criteria, submission cohort stratification, and final matched cohort stratification, EHR = Electronic Health Record.

| **Description** | **N** | **% of patient population** | **% of submission** | **% of matches** |
| --- | --- | --- | --- | --- |
| ***Total patients meeting inclusion criteria*** | ***307,799*** | ***100%*** |  |  |
| *Recorded death in EHR* | 29233 | 9.5% |  |  |
| *Unknown status in EHR* | 278576 | 90.5% |  |  |
| ***Total Submitted to NDI*** | ***35015*** | ***11.4%*** | ***100%*** |  |
| Recorded death in EHR | 12395 | 4.0% | 35.4% |  |
| Unknown Status in EHR | 22620 | 7.3% | 64.6% |  |
| **Final Matches** | ***13709*** | ***4.5%*** | ***39.2%*** | ***100%*** |
| Recorded death in EHR | 11,934 | 3.9% | 34.1% | 87.1% |
| Deceased with no recorded death in EHR | 1775 | 0.6% | 5.1% | 12.9% |

*EHR = Electronic Health Record*

### 3.2 Summary of linkage of IIM Records to EHR Records

The initial probability cutoff set within the probabilistic matching algorithm was 0.80 which resulted in the vast majority of the reference cohort having a potential match identified. The highest probability match for each patient within the reference cohort was assessed. We subset these based on match probability and assessed each of these grouping for match validity to determine final match criteria (Table 2). Exact matches on first name, last name, and date of birth were identified for 37.2% of the MGB cohort and 46.5% of the VUMC cohort.

**Table 2:** Matching between EHR and Social Media Records using a probabilistic linkage algorithm, MGB = Mass General Brigham, VUMC= Vanderbilt University Medical Center.

| Description | MGB Data |  |  | VUMC Data |  |  |
| --- | --- | --- | --- | --- | --- | --- |
|  | N | % of matched | % of total MGB death cohort | N | % of matched | % of total VUMC death cohort |
| Total death in MGB reference data | 65,139 | - | - | 13,709 | - | - |
| Total deaths in publicly available data | 8,132,598 | - | - | 8,132,598 | - | - |
| Total patients matched with $p \geq 0.80$ | 56,485 | 100% | 86.7% | 13343 | 100% | 97.3% |
| *Match Probabilities* |  |  |  |  |  |  |
| <0.975 match probability | 20,281 | 35.9% | 31.1% | 12 | .09% | .09% |
| $\geq 0.975$, not exact match | 11,976 | 21.2% | 18.4% | 6952 | 52.1% | 50.7% |
| Exact match | 24,228 | 42.9% | 37.2% | 6379 | 47.8% | 46.5% |

MGB = Mass General Brigham, VUMC= Vanderbilt University Medical Center

### 3.3 Validation results for linkage of IM Records to EHR Records

Potential matches stratified by probability were manually reviewed to determine if they were likely to be a true match with an emphasis on reviewing matches ranked as non-exact matches. Both institutions found that matches with high probability (>.99998) matched on names and DOB were deemed a “true” match, but that matches with lower probability only met the criteria in our review for a “true” match in less than 50% of the reviewed pairs (Tables 3). This is even with a broad definition of match which only required first name match and DOB within 5 days or an uncommon full name match. The results indicated that to reduce the level of false positive matches, the match criteria needed to require a full name and DOB match between IM and the EHR.

**Table 3:** Validation of probabilistic linkage of Internet Media Records to EHR Records by manual review DOB = Date of birth, DOD = Date of death.

| Match probabilities | Total records | Records manually reviewed | Confirmed matches by manual review | % Accuracy | Observations from manual annotation |
| --- | --- | --- | --- | --- | --- |
| **MGB data** |  |  |  |  |  |
| <0.975 | 20,281 | 200 | 89 | 45% | In most matches, two out of four fields matched (first & last name; first name & DOB; last name & DOB) |
| >=0.975, not exact | 11,976 | 32 | 15 | 47% | Most matched exactly on first name and last name + some matched closely on DOB (within 30 days) and state |
| Exact | 24,228 | 68 | 68 | 100% | Matched exactly on first name, last name, DOB, and state |
| Total (>=0.8) | 56,485 | 300 | 172 | 57% | - |
| **VUMC data** |  |  |  |  |  |
| <0.975 | 12 | 12 | 12 | 100% | Matched on first name and date of birth, mismatched on last name and state. DOD did not match reference. |
| >=0.975, not exact | 6952 | 225 | 13 | 5.7% | Most matched first name and last name but mismatch on DOB with some matches on state. The accepted matches matched on last name and DOB with variant first names (nickname/typo). |
| Exact | 6379 | 63 | 63 | 100% | Matched on first name, last name, DOB, and state |
| Total (>=0.8) | 13343 | 300 | 88 | 29.3% |  |

DOB = Date of birth, DOD = Date of death

We further reported PPVs by probability groups based on alignment of dates of death (Table 4). Predicted matches from the probabilistic matching procedure that matched exactly on the first name, last name, and date of birth showed PPV of 94-99%. Predicted matches that did not exactly match on these variables showed PPV of <6%, suggesting that these matches were likely inaccurate matches.

**Table 4:** Positive predicted values for deaths identified with internet media records based on alignment of dates of death of reference standard.

| **Match probabilities** | **Total records** | **True positives based on dates of death within 7 days of the reference standard** | **Positive predicted Value (death date within 7 days)** | **True positives based on same date of death as reference standard** | **Positive predicted Value (same death date)** |
| --- | --- | --- | --- | --- | --- |
| **MGB data** |  |  |  |  |  |
| <0.975 | 20,281 | 121 | 0.5% | 23 | 0.1% |
| >=0.975, not exact | 11,976 | 312 | 2.6% | 243 | 2.0% |
| Exact | 24,228 | 23,802 | 98.2% | 22,979 | 94.7% |
| **VUMC data** |  |  |  |  |  |
| <0.975 | 12 | 0 | 0.0% | 0 | 0.0% |
| >=0.975, not exact | 6,952 | 382 | 5.6% | 347 | 5.1% |
| Exact | 6,379 | 6,301 | 98.9% | 6,135 | 96.3% |

### 3.4 Augmentation and Evaluation of Death Data

#### 3.4.1 Augmentation of Death Data at MGB

Among the 65,389 deaths in the MGB reference cohort, the sensitivity of capture was 33% for EHR alone (n=21,595) and 37% for IM records (n= 24,228) when using exact match. Overall, IM records using exact matching identified an additional 15,661 deaths compared to the EHR alone, increasing the sensitivity from 33% to 57% (Figure 2).

#### 3.4.2 Augmentation of Death Data at VUMC

Within our reference cohort of 13,709 patient, 6,379 (46.5%) had an exact match within IM sources. Of 11,934 with recorded death dates, 5,629 (47.2%) had IM records and of the 1,775 with unknown death status in the EHR, 750 (42.3%) had a record in IM sources (Supplementary Figures 1A, B). Given that our submission was not a random representative sample, we needed to extrapolate the actual rates of death in our cohort population based on the NDI results and from that calculate the rates of recorded deaths and the rate of augmentation from IM sources. As mentioned, the sample VUMC submitted to NDI to obtain our reference standard was skewed for known deaths with 35% of the total submission having recorded dates of death within the EHR. Based on a rate of 7.8% deaths within our submitted unknown sample and the numbers of patients with unknown death status within the full 2019-2021 VUMC cohort, we can estimate an additional 21,729 death over that time for a total of 50,952 deaths (Supplementary Figure 1C). Using that as the basis for calculation, we estimate that 57.3% of the total 2019-2021 VUMC deaths are captured within the EHR. In our reference data, IM death data existed for 42.3% of deceased patients which would extrapolate to an additional 9191 deaths, an increase of 18.0% (Figure 3B), comparable to the 24% increase seen at MGB.

## 4.0 Discussion

In this validation study conducted across two large healthcare systems from the US, we observed substantial missing data in the recording of deaths within EHRs. We further observed that leveraging NLP models to extract death information from publicly available IM platforms can accurately identify an additional 18-24% of deaths not captured in EHRs.

Mortality is a key outcome in observational studies, making capture of accurate death information of high importance. Many observational research studies rely upon data collected during the course of standard clinical care and available within a subject’s EHR record, but these sources have missing mortality information^3, 9^. Missing mortality information consistently leads to overestimate of survival with increasing levels of missingness relating to an increased bias in survival estimates^3, 5, 34^. Even low levels of missing data potentially have significant impacts when assessing long-term survival and prevalence^35^. Low sensitivity of mortality data must be addressed in order for the data to be considered fit for purpose ^4, 5, 8, 36^.

The standard sources for death data outside of the EHR include state vital statistics departments and NDI. NDI is often considered a “gold-standard” source for death information, but can face about 2 years lag before death data are made available^18^. For state vital statistics, depending on the study question, investigators may need to petition for access to multiple state registries and some gaps in coverage may still exist if patients move between states. For these reasons, death information extracted using NLP from publicly available internet sources such as obituaries, memorial archives, such as Everloved/TributeArchives, and other social media sharing sites, like GoFundMe, may be a useful supplement to EHR recorded death information. To support others in utilizing these models for similar purposes, we have made our NLP models publicly available for future use^37^.

Prior studies have attempted to use IM data to extract fact of death, but these have primarily focused on obituary data in small cohorts^10, 11, 29^. Our study expands on the literature by providing by far the largest validation of death information extracted from online sources including obituaries, memorial websites, and other social media, against gold standard information from state vital files or NDI. Importantly, our findings are likely more generalizable than prior studies as we conducted a population-based validation using deaths from two large healthcare systems in the US without restricting to any specific subpopulation with specific conditions.

There are some caveats and limitations to the use of IM sources for death ascertainment that merit discussion. Our results indicate that a high level of stringency needs to be used in determination of matches to avoid false positives and erroneous assignment of deaths, which is in line with prior work^29, 38^. We further acknowledge that even after successfully extracting the information from social media and linking them with EHRs, the sensitivity of mortality capture remains suboptimal. Finally, there is little literature on possible bias when using online memorial websites and obituaries for research purposes, but the possibility of non-representativeness in the sample with deaths recorded in internet media sources cannot be ruled out given the cost-related barriers associated with publishing obituaries and reliable internet access.

Overall, IM sources have the potential of increasing coverage of mortality outcomes data by 18-24% over reliance on EHR records alone with the ability to obtain this information in near real-time. For timely real-world surveillance, IM sources offer a potential solution to capture mortality information without the need for release of any PHI data outside of institutional firewalls, as public IM data can be scraped and brought into a secure environment for matching with EHRs.

## Data Availability

All data produced in the present study are available upon reasonable request to the authors

---

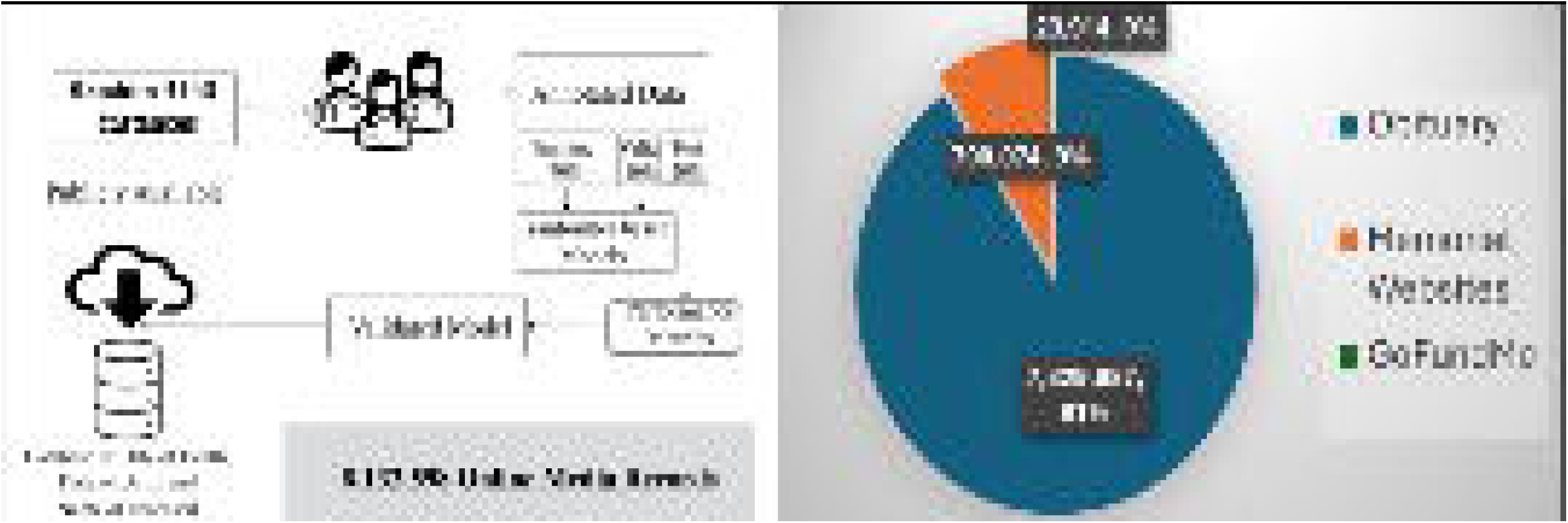

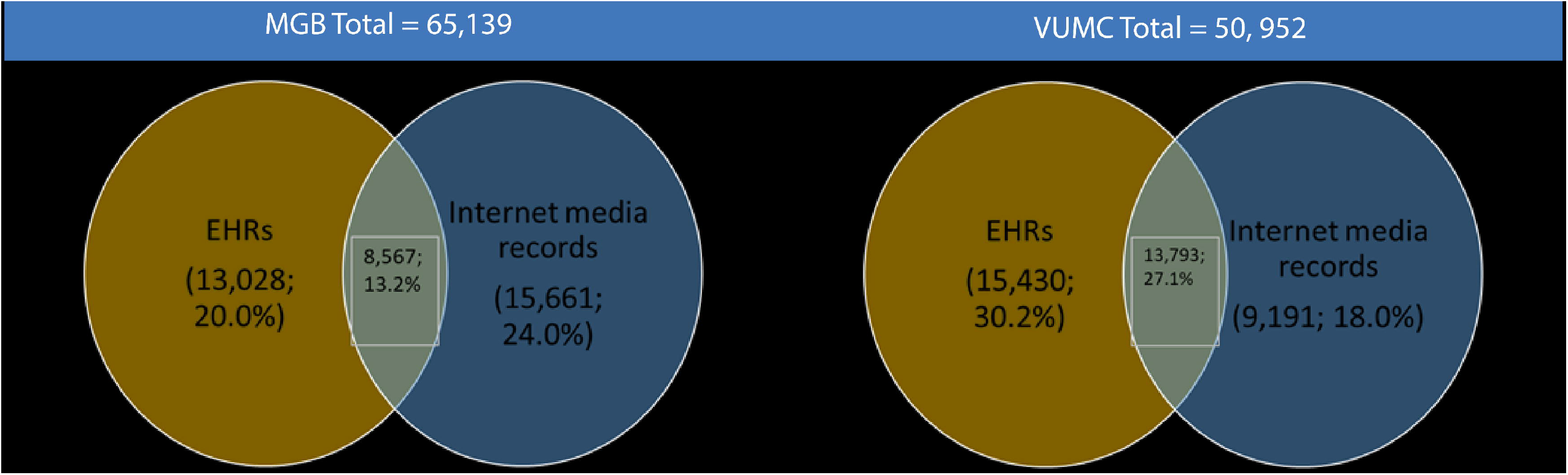

