## Supplementary material for "Augmenting Fact and Date of Death in Electronic Health Records using Internet Media Sources: A Validation Study from Two Large Healthcare Systems": Figure captions

***Figure 1: Public Internet Media Data.*** *A) Internet Media sources data extraction methods B) Distribution of records from specific sources*

***Figure 2: Augmentation of death data through addition of internet media recorded deaths to electronic health records (EHRs)****.*

*NOTE: For VUMC (on right), the Venn diagram shows number of deaths recorded in EHR (2019-2021) and estimated deaths that would be identified in internet media records given rates of identification from reference cohort.*
