## Supplementary figures for "Augmenting Fact and Date of Death in Electronic Health Records using Internet Media Sources: A Validation Study from Two Large Healthcare Systems"

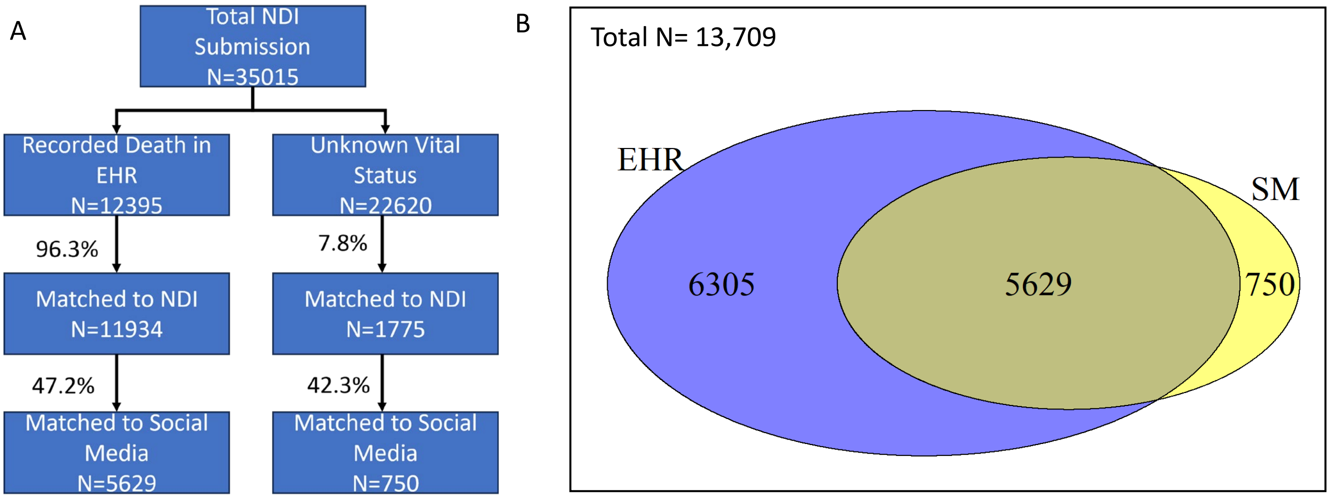


C

| Year | Recorded Death Date | No Recorded Death Date | Estimated Unrecorded Deaths |
|---|---|---|---|
| 2019 | 8,995 | 89,120 | 6,951 |
| 2020 | 10,597 | 94,188 | 7,347 |
| 2021 | 9,631 | 95,268 | 7,431 |
| **TOTAL** | **29,223** | **278,576** | **21,729** |

**Supplementary Figure 1: Internet Media Augmentation.** A) Diagram showing match rate from EHR submission to NDI, breakdown of recorded and unrecorded deaths in EHR within the reference cohort, and linkage with public online data. B) Venn diagram showing number of deaths recorded in EHR (blue) and number in social media (yellow), C) Estimated deaths within VUMC patient population from 2019-2021 given 7.8% rate of death with unknown vital status cohort
